# Healthcare workers’ mental health during the COVID-19 pandemic: A qualitative analysis of a text message-based NHS workforce support line

**DOI:** 10.1101/2024.08.16.24311723

**Authors:** Lisa J. Gould, Eleanor Angwin, Richard A. Powell, Emma L. Lawrance

## Abstract

**Background:** The National Health Service (NHS) is suffering from a workforce crisis of mental and physical sickness and attrition following the COVID-19 pandemic. An in-depth understanding of healthcare workers’ (HCWs) experiences during the pandemic is required to understand the impacts on their mental health in this challenging work environment. This qualitative study explores HCWs’ concerns during the COVID-19 pandemic - expressed in real-time during an active mental health crisis.

**Design:** This study involved analysis of data from “Shout”, a text message-based, UK-wide mental health support service which, during the pandemic, was advertised to HCWs specifically. Pseudo-random sampling of scripts of anonymised text message conversations between HCWs and Shout Volunteers from April 2020 – March 2021 was undertaken, with data fully anonymised by Shout before researchers accessed them on a secure purpose-built platform. Following application of exclusion and inclusion criteria, 60 conversations were coded to develop a thematic framework and analysed using grounded theory, with sub-themes triangulated to create final themes. Quotes extracted from this process were then synthesised for publication.

**Results:** Three themes emerged from the data: 1) Poor mental health, sub-themes: (a) overwhelming negative feelings or emotional distress experienced, and; (b) active crisis/resurgent symptoms. 2) Negative work experiences, sub-themes: (a) negative NHS work culture and expectations; (b) inadequate structures and arrangements for support; (c) trauma at work, and; (d) abuse at work. 3) The impact of the COVID-19 pandemic, sub-themes: (a) additional work pressure, and; (b) isolation and risk.

**Conclusion:** This study explores the challenges and mental health concerns in HCWs during an active crisis. Organisational stressors, mental health provision and additional resources for HCWs to recover from the pandemic remain a vital issue in current NHS service provision.

**Strengths and limitations:**

- This study uniquely provides an analysis of HCWs’ mental health experience, recorded in real-time across the pandemic, thereby free from recall bias.
- The study analysed insights from a unique and valuable dataset of particularly vulnerable HCWs (i.e. those in active crisis) in their own words, enabling rich insight into the nature and circumstances of severe distress in HCWs.
- The individuals using the Shout text service are more likely to be in a severe mental health crisis and not therefore representative of the whole HCW community, but given the study is aligned with the principles of qualitative enquiry, we do not seek to be representative of, or generalizable to, the whole population.
- This paper is not able to represent the views of all HCW’s, and whilst we consider different groups of HCW’s in the body of the analysis, there are relatively small numbers in each group.

## Introduction

The National Health Service (NHS) is experiencing “the greatest workforce crisis in their history” (1); NHS trusts have lost more than 560,000 days due to staff anxiety, stress, depression or another psychiatric illness(1). HCWs are known to be at risk for anxiety, depression, burnout, insomnia, moral distress (i.e., an inability to do the ‘right thing’), and post-traumatic stress disorder (2–5). This effect was exacerbated by COVID-19 (6–12), with the United Kingdom (UK) and global communities struggling to create resilient healthcare systems (13,14). Recently released NHS long-term plans to address this emergency include a commitment to “support the health and wellbeing of [the] … workforce” (15).

In 2020 *The Lancet* published a communication considering how NHS staffs’ mental health could be supported during the pandemic (16). The report summarised the mental and physical challenges staff faced and stressed the importance of post-trauma support for NHS staff, noting that anonymous mental health tools might help. During the pandemic, organisational interventions were initiated to support NHS workers’ mental health, including targeted advertisement of the “Shout” service to HCWs (https://giveusaShout.org/get-help/our-frontline-you-support-us-we-support-you/). Shout is a free, anonymous, text message-based mental health support line available to anyone in the UK. It is targeted at people feeling overwhelmed and actively distressed and provides support via trained volunteers supervised by clinicians on a specialised platform.

Research shows digital and mobile interventions can have positive benefits for staff health and wellbeing (17,18) and improve service provision to population subgroups not typically using health services (19). However, despite evidence of their success and frequency of use— with Shout hosting 2.5 million mental health conversations to date—and with both the public and HCWs, such support services are experiencing depleted funding and closures. The Royal College of Nursing, for example, recently called for urgent psychological support for nurses, voicing concerns regarding the potential closure of further specialist mental health hubs. The British Medical Association have also instigated calls for better mental health support for doctors and General Practitioners (GPs), citing a 2022 survey which found almost a quarter of GPs knew a colleague who had died by suicide (20) and noting calls from GPs to its counselling service more than tripled during 2021-2022(20).

To better understand the stressors driving continued mental health support for NHS workers post-pandemic, there is a clear need to better understand the experiences of HCWs affected by a mental health crisis or distress during the pandemic. In particular, there is a need to ascertain the contributing factors to worsening mental health and active crisis, and identify what support is helpful and where it should be targeted. A deeper understanding of factors exacerbating mental distress can offer targets for healthcare system improvement to benefit staff and ultimately patient safety (21,22). Furthermore, understanding who, how and why HCWs engage with crisis support services can guide appropriate service provision to safeguard HCWs experiencing distress.

This paper therefore explores anonymised data from the Shout service. In April 2020, Shout launched their ‘Our Frontline’ programme, working with the NHS and other charities to support key workers through the pandemic (23). In 2022 a small group of trained researchers were given access to a subset of fully anonymised conversations from Shout on a specifically-designed highly secure online platform. This study aimed to understand the experiences of UK HCWs who used the Shout text messaging service over the first year of the COVID-19 pandemic through anonymised text-based conversations between the service and UK HCWs.

## Methods

### Study design

This study is a thematic analysis of a subset of qualitative data of fully anonymised text message conversations from the Shout text messaging service.

### Patient and public involvement

None. This research was conducted mid pandemic, and we were unable to engage this population at the time. Due to the sensitive nature of the data used, legal clearance was required from MIH to make the details of this study public, to anyone including PPI.

### Data collection

#### Data sources

The dataset consisted of fully anonymised text message conversations between HCW texters and Volunteers from the Shout service in the 12-month period from April 2020 to March 2021. The dataset owned by the Shout service and was made available to highly trained researchers on a bespoke highly-secure online research platform.

#### Data Extraction/Screening

Initial screening of the text message-based dataset was conducted by Shout, which selected 3,476 conversations using the relevant texter keyword FRONTLINE corresponding to the “Our Frontline” conversations.

A cut-off point was used to include only conversations of sufficient length to provide enough richness and detail for the qualitative analysis. After consultation with the Shout team the researchers used conversations over 35 messages in length; all shorter conversations were excluded, leaving 1522 conversations (Table 1). The research team estimated 60 conversations would provide saturation for a thematic analysis-the unique design of this study resulted in a scarcity of previous similar studies to draw upon, so this estimate was based on previous experience and length of text message transcripts. Conversations were therefore pseudo-randomly selected from the 1522 conversations and screened using inclusion and exclusion criteria (Table 2); they had to include a reference to HCW status to be included and were excluded if they contained a statement that negated HCW status. This process was continued iteratively until 60 appropriate conversations had been selected, 96 conversations in total were screened, and 36 were rejected (Table 1).

**Table 1:** Selection process of anonymised conversations showing how many conversations were included or excluded.

| Total conversation | Screening | Number Included | Number Excluded |
| --- | --- | --- | --- |
| 3,476 conversations | Screened for $\geq 35$ messages<br>in total | 1522 included | 1954 excluded |
|  | Randomly selected conversations, screened further for active reference to HCW status or statement that negates HCW status. | 96 included | 36 excluded |
|  | Final conversations randomly selected to be included in analysis | 60 conversations |  |

**Table 2.** Summary of inclusion and exclusion criteria for purposive sampling of anonymised conversations with healthcare workers from the Shout dataset.

| Inclusion criteria | Exclusion criteria |
| --- | --- |
| • $\geq 35$ messages in total<br>• Active reference to HCW status (e.g. I am a nurse). | • $\leq 35$ messages in total<br>• Statement that negates HCW status i.e. social care worker. |

### Data analysis

Two coders were involved in this study (L.G, E.A). Both coders were White British females, educated to postgraduate level, with extensive experience working in healthcare systems, one as a manager and the other as an academic. The researchers align with an interpretivist epistemology (24). A grounded theory approach was taken to enable the generation and development of themes (25). The qualitative analysis software NVivo 12 was used to analyse the anonymised conversations. The analysis occurred in three steps. First, a single coder (E.A) used open coding to code text line by line to minimise preconceptions and enable comparisons between each texter’s conversation. Second, axial coding was used to identify relationships between the codes and develop categories. Finally, selective coding was used to connect all categories into themes and sub-themes. The final themes were co-developed with the principal investigators (L.G and E.L), and a peer researcher support group was used to corroborate emerging findings (26). The coding tree is illustrated in Figure 1.

**Figure 1:**
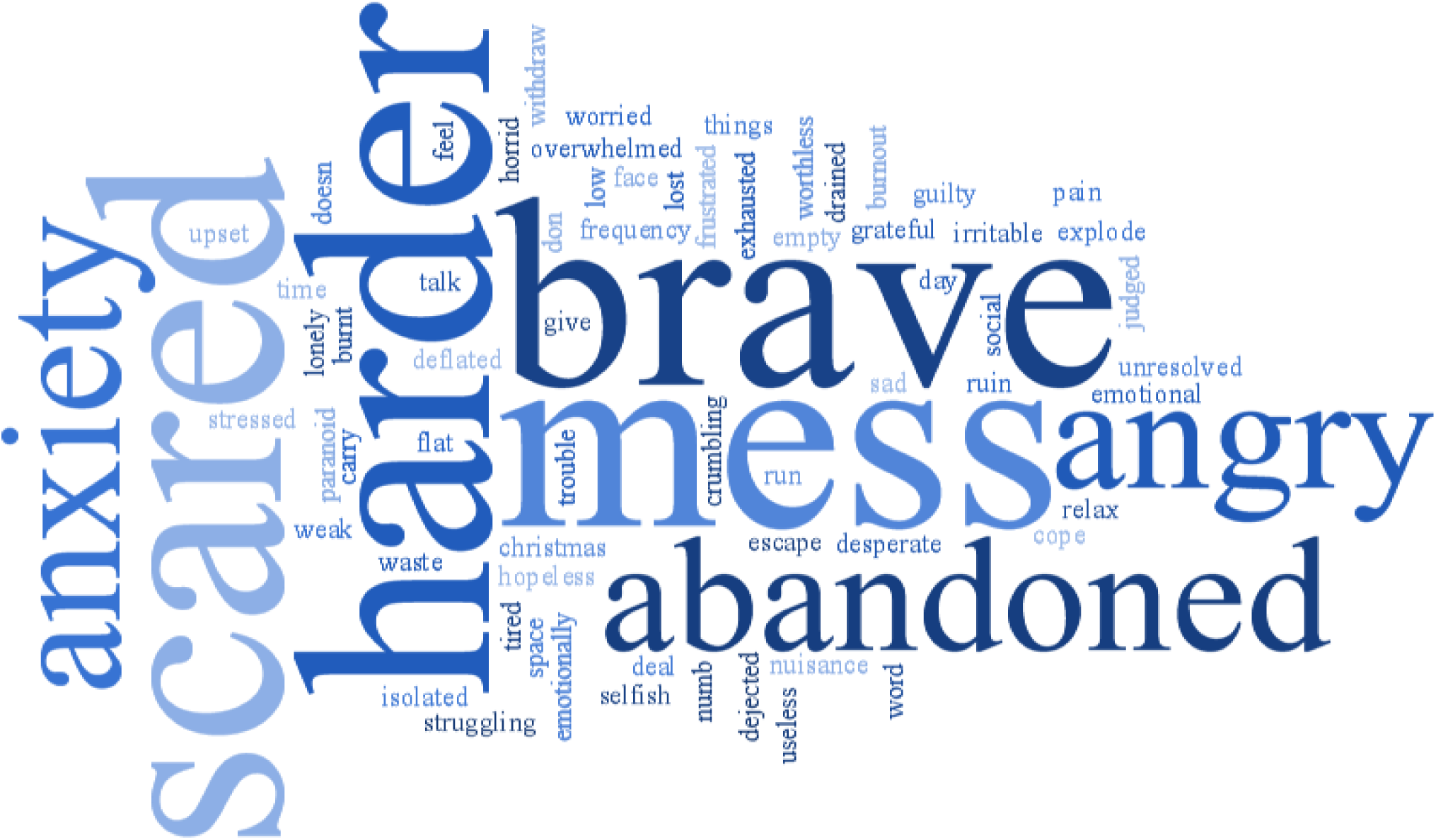
Coding tree used to generate themes

**Figure 2:**
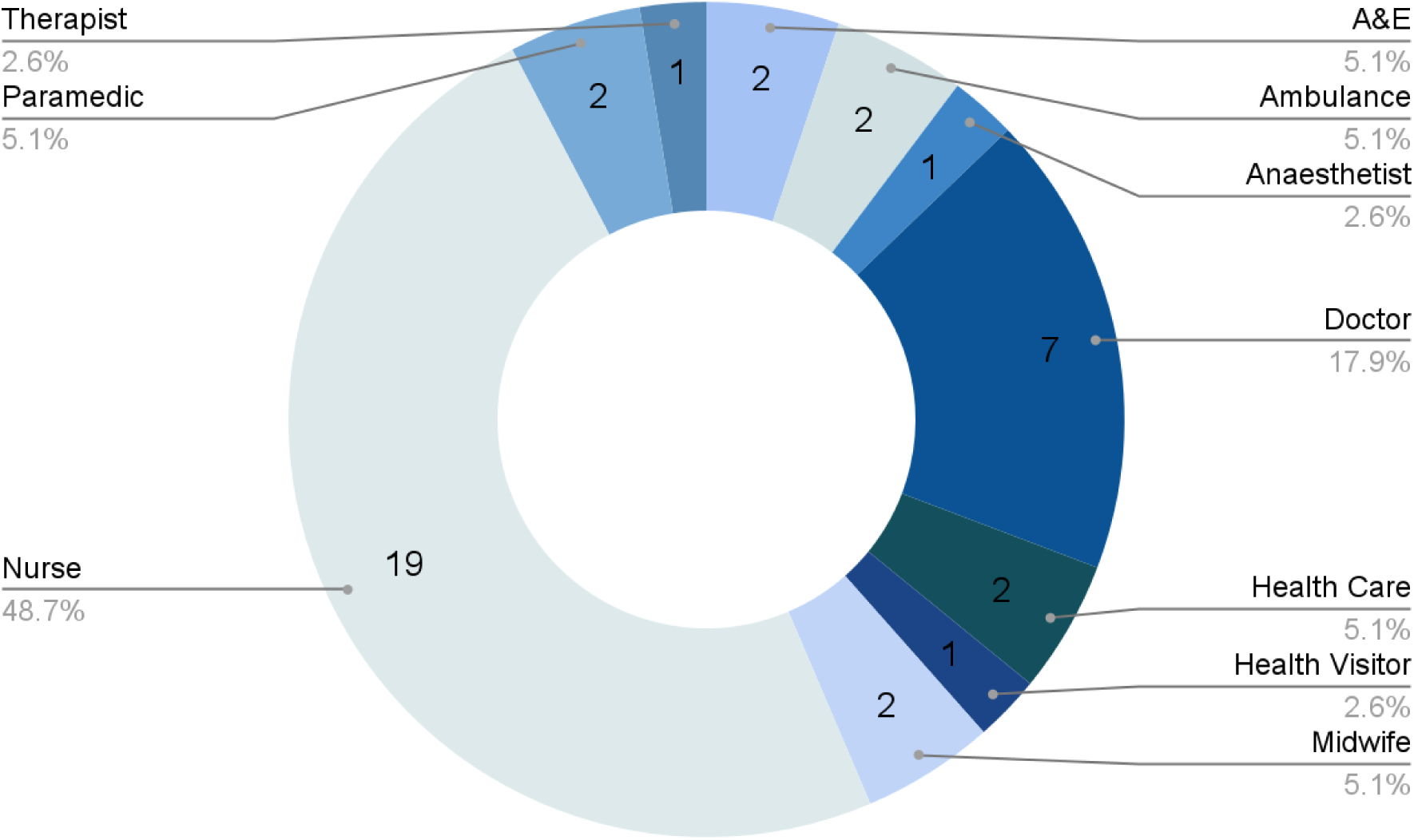
Distribution of the type of HCW who identified their professions

## Results

### Caller professions

In addition to the FRONTLINE keyword, many HCWs explicitly referenced their profession during conversations. Nurses were the most common HCW in the sample of conversations to self-identify professionally when contacting the service, with doctors second.

### HCWs’ mental health status

There were three self-identified mental health statuses used by those contacting the service in the sample of conversations. First, HCWs experiencing an acute crisis; for example, an ongoing panic attack or plans in place for self-harm or suicide. Second, HCWs identifying themselves as having pre-existing mental health conditions, which they felt had worsened over the pandemic. Third, HCWs dealing with emerging symptoms or experiences of mental health conditions. The most common self-identified mental health conditions or symptoms of HCWs texting into the service were depression and anxiety, with the majority of texters who identified themselves as having pre-existing conditions either having had a previous diagnosis of, or experience with, depression or anxiety.

Doctors in our sample (noting this is a minority of conversations) were more likely than other types of HCW to be experiencing severe distress; i.e., more at risk of imminent or past self-harm, or a high-risk suicide case. Nurses, on the other hand, tended to predominantly report experiencing depression and anxiety.

### Thematic findings

Three primary themes (poor mental health, negative work experiences, and the impact of COVID-19), and eight subthemes, emerged from the data. Synthetic quotes (where the quotation was altered slightly by the use of synonyms or minor changes to context) were used to enable the illustration of the themes whilst maintaining texter anonymity. The composite “quotations” used were chosen as illustrative of the key themes from the paper - either through being commonly voiced or through highlighting a particular viewpoint shared by certain individuals, including those representing minority or dissonant but nevertheless important themes.

### Theme One: Poor mental health

#### (a) Overwhelming negative feelings or emotional distress experienced

HCWs’ experiences of poor mental health focused predominantly on symptoms consistent with burnout, anxiety and depression. They directly identified their symptoms as manifestations of anxiety, depression and burnout, or indirectly referenced symptoms that can be indicative of these mental health challenges, through the use of words such as drained, exhausted, flat, overwhelmed, stressed, dejected, useless, and desperate. See Figure 3 for a word map of the emotive words used most frequently by texters in the sample.

**Figure 3:**
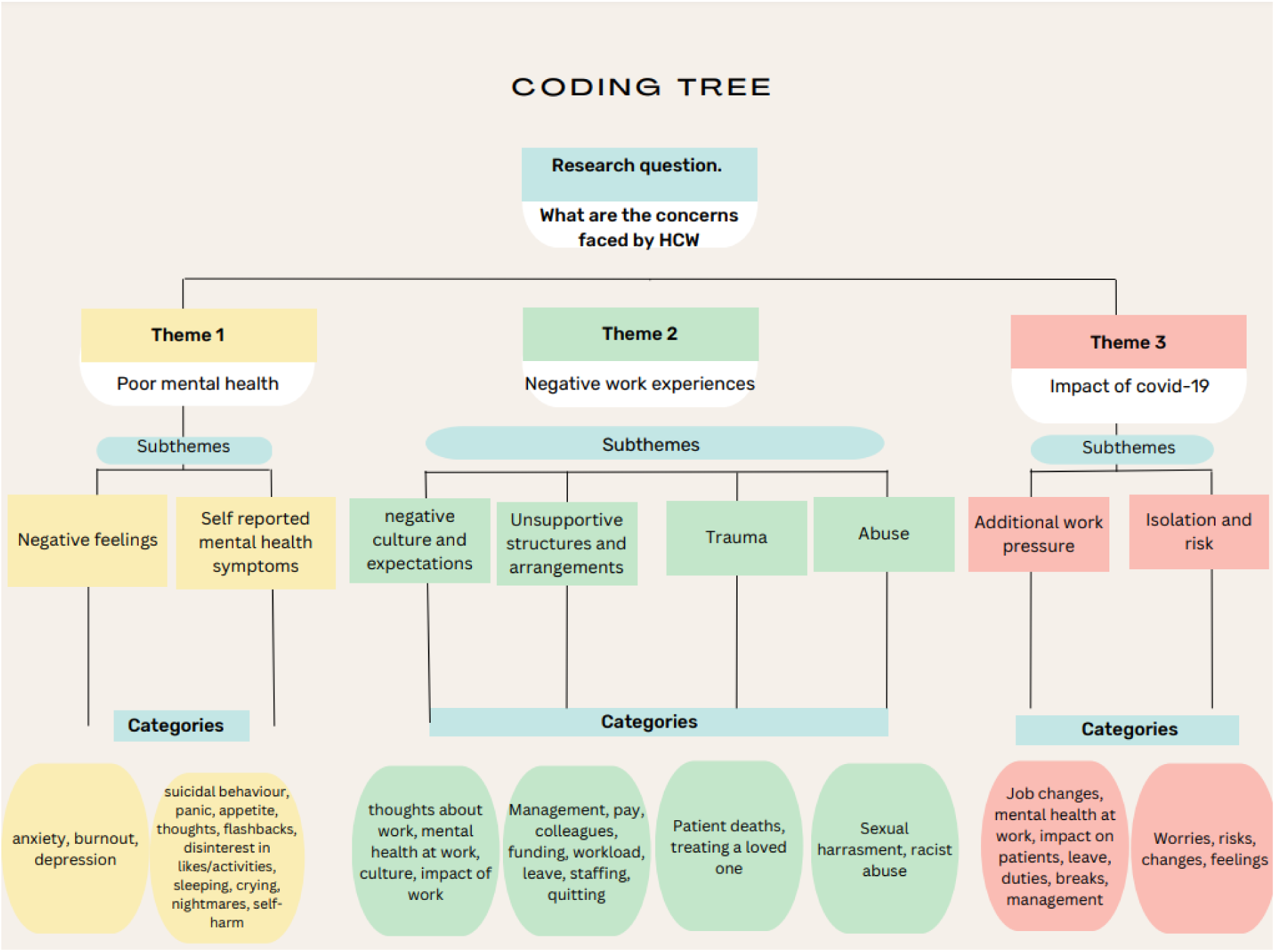
Word map of emotive word use

#### (b) Active crisis/Resurgent symptoms

A large proportion of HCWs texting into the service in our sample were also experiencing an acute mental health crisis, either through experiencing a current panic attack, or feeling suicidal and being at high-risk due to having a plan in place and having access to means. Many reported experiencing resurgent symptoms of mental health conditions, most commonly difficulty sleeping. Of the doctors included in this sample, most appeared to experience more severe mental health crises, self-harm, suicidal ideation and mental breakdowns.

### Theme Two: Negative work experiences

Work experiences had a tangible impact on individuals’ mental health, with work often being an explicit cause or trigger of symptoms, including intrusive recurring thoughts (most commonly reported in sample as symptom linked to work), panic attacks, suicidal ideation or a full mental breakdown. HCWs experienced these symptoms at home, but most commonly reported experiencing them directly before, at, or after work.

#### (a) Negative NHS work culture and expectations

Fear and bullying were highlighted as negative experiences when describing the culture of the NHS and being a HCW. Texters reported feeling fear; they felt unable to make mistakes in their roles or ask for help without fearing “consequences” from management. Experiences of bullying described by HCWs included inappropriate behaviour by both management and peers. In these instances, HCWs felt like there was no accountability for those who behaved poorly.

*“I cannot stand the NHS anymore. They don’t care about us and we can’t raise it as managers think it’s just an excuse not to work.”(synthetic example quote)*.

Whilst some texters continued to feel positively about their jobs, the majority could only describe their job negatively. Several were either considering leaving their jobs or reported seeing an increase in colleagues leaving over the last year. A key sentiment of those who felt positively about their jobs was that they really cared about the work they did. Those who felt negatively often reflected sentiments of burnout.

Many HCWs, particularly the doctors and nurses included in this sample saw their profession as a key feature of their identity. Nurses tended to view the caring duties associated with their role as inherent to their nature. This could often come at the expense of their own wellbeing.

*“We [as nurses] are expected to look after others, all the time, even if it means that we are unable to look after ourselves.”(synthetic example quote)*

The doctors in this sample tended to view their profession as synonymous with being strong. If they were exhibiting mental health or emotional wellbeing symptoms (e.g., crying), they tended to view this as being weak.

*“As a doctor. I know that I need to be stronger than this - I don’t understand why I am feeling this weak.” (synthetic example quote)*

In both their work and home environment, there was a reported culture of needing to put on a brave face and ‘power through’ with their jobs despite struggling. Often HCWs described needing to meet expectations put upon them, either because of their beliefs about their profession as part of their identity, or because of expectations from others. In several instances, HCWs were told by Shout volunteers how brave they were for doing their jobs - this was almost always met with HCWs reporting feeling they did not match those expectations. Additionally, in one instance, the “Clap for Carers” (i.e., weekly applause given by the public for HCWs during the pandemic) was mentioned as a direct contributor to poor mental health as it perpetuated a feeling of needing to put on a brave face.

#### (b) Inadequate structures and arrangements for support

##### Unsupportive management and colleagues

For the majority of HCWs texting in who were concerned about experiences at work, strong negative experiences were related to work environment structures and arrangements. Most described feeling unsupported by managers and peers, manifesting in feeling unable to ask for help or share their feelings with those at work due to negative relationships, lack of support, or not wanting to burden others who they could see were already stretched. Where positive management relationships were described, it often manifested in management listening to their concerns, being approachable and making adjustments for the HCW to support them (e.g., changes to shift patterns). Unsupportive management was most often characterised as uncaring, behaving inappropriately, providing unrealistic pressure, and not checking in on staff members.

*“My manager doesn’t care about me. They told me I needed to return because I’d taken too much time off – I’d been off for two days for a family bereavement ” (synthetic example quote)*

While relationships with peer colleagues were more often reported to be positive than relationships with management, there was still a powerful sentiment of feeling unsupported by peer colleagues. For some HCWs, this manifested in inappropriate behaviour or feeling a sense of difficulty in getting on with colleagues. For the majority of HCWs, the unsupportive nature of colleagues stemmed not from an active intent, but instead from pressures and stresses faced by the overall team that meant colleagues were disengaged or unavailable to support each other.

*“I don’t really have supportive colleagues - everyone is so stressed I don’t feel able to talk to them about any of this.” (synthetic example quote)*

##### Unable to take time off

A consistent theme in HCWs’ experiences at work was an inability to take time off and take time for themselves. Many HCWs struggling with their mental health felt unable to take sick leave. This was due to either being fearful of the consequences with, and perceptions of, management or, more commonly, not wanting to let their team down. In instances where individuals had been signed off sick and were due to return to work, there were strong fears around whether they would be able to cope and also instances of senior management pushing for HCWs to return to work from sick leave before they were ready.

Shout *service provider: “Are you able to take time off? It is important to take care of yourself.”*

*HCW: “I can’t. I know I need to but my manager will be really unhappy with me if I do. They really don’t like people taking time off sick.” (synthetic example quote)*

##### Increased workload

Many HCWs described an increase in their workload and work intensity as causing additional concerns. Work was consistently described as getting busier, with HCWs working longer hours, more shifts, and seeing more patients due to the pandemic. Additionally, a large number of HCWs had seen a change in their roles and were given additional high-risk responsibilities with minimal training or clarification of expectations of their roles, leaving them struggling to cope. Many felt unable to complain or highlight their concerns to management.

*“Work is so busy and I feel like what I’m being asked to do is dangerous. I don’t feel like I can speak to my manager about it - I’m frightened they’ll think I’m just trying to use excuses to get out of work.” (synthetic example quote)*

#### (c) Trauma at work

HCWs were experiencing traumatic incidents at work which stuck with them and impacted their mental health. In a large number of cases, the loss of a patient was a strong contributor to HCWs’ poor mental health or distress. Often the unexpected or traumatic loss of a patient led to recurring intrusive thoughts for the HCW. In several of the suicidal cases, doctors were blaming themselves for either failing to save a patient or having to diagnose a patient with a terminal illness. In these instances, HCWs felt significant guilt and that they no longer deserved to live given they could not save their patient. There were also multiple occasions where a HCW’s loved ones were admitted to their place of work with COVID-19, leaving them with recurring intrusive thoughts that impacted their ability to do their job both while their loved ones were at their place of work and afterwards.

*“I couldn’t save them. I do not deserve to live.” (synthetic example quote)*

Work also became a trigger for re-experiencing trauma.

*“I keep having nightmares that I’m going to have to go back to the Intensive Care Units [ICUs].” (synthetic example quote)*

#### (d) Abuse at work

There were also several incidents of abuse towards the HCWs themselves. There were multiple instances of harassment on the job, including racial and sexual harassment by patients. These incidents left HCWs feeling unable to perform their duties adequately, which they viewed as a risk to their patients.

*“I don’t feel comfortable being alone with patients now after what happened.” (synthetic example quote)*

### Theme 3: Impact of COVID-19 pandemic

COVID-19 provided an additional burden at work and in HCWs’ personal lives.

#### (a) Additional work pressure

Work pressures were exacerbated by COVID-19 for the majority of HCWs, who saw an increase in patients dying or becoming sick. Several described having to make difficult decisions about prioritising patient treatment and beds. A common experience of HCWs working on COVID-19 was they were redeployed to new roles, most often in ICUs. Often these moves came without any support from management either before, during or after the redeployment, where the expectation was for them to just return to their teams. For those HCWs who did not work explicitly on COVID-19 wards, their work experiences were still impacted by the pandemic through cancelled learning and development or annual leave.

*“I was moved to another team during COVID – not once did my manager check in with me over months. Now I’m expected to come back to my team and have a relationship with someone who didn’t care about me at all.” (synthetic example quote)*

#### (b) Isolation and risk

The pandemic also impacted HCWs’ personal lives. The majority of HCWs experienced concern about the additional risks that came with their job, fearing mostly they would infect loved ones or vulnerable patients.

*“They want me back doing visits. I’m frightened of infecting my family – they are vulnerable to COVID.”(synthetic example quote)*

Many were also worried for their own health, either in the abstract of a fear of dying, or in specific circumstances where they were being either forced or felt they had to work despite being high-risk or shielding. Isolation was a powerful experience of HCWs during the pandemic, with many feeling their inability to see loved ones or do things they would usually enjoy was worsening their mental health.

## Discussion

Thematic analysis of anonymised conversations between HCWs and a text message-based support service across the first year of the COVID-19 pandemic identified three main themes characterising their experiences and contributing factors. These were (a) poor mental health; (b) negative work experiences; and (c) the impact of the COVID-19 pandemic. The experiences shared by texters using this service gives a real-time and raw account of the severity of mental health problems experienced by NHS workers, which appeared in many incidences to be directly related to their professional experiences. There was evidence of distress caused by bullying and fear culture, unrealistic or misaligned expectations from the public, pragmatic managerial issues, trauma, abuse and the pressures of the pandemic.

This study identified particular risk factors for HCWs in the context of the COVID-19 pandemic, with HCWs self-identifying as having prior mental health conditions potentially particularly vulnerable to mental health crisis and associated harmful behaviours. Our study also indicated that doctors, who are traditionally less likely to respond to wellbeing surveys (27), could also be a high-risk group. From previous literature, evidence suggests doctors are prone to mental health stigma due to culture and their work environment, and its perceived negative professional impact (28–30). This, combined with doctors being less likely to register for, or access, GP services, and research corroborating doctors mental health experiences during the pandemic (21,31) may explain why doctors present the most severe cases seen by the Shout service in our sample (28,29).

This study adhered to guidance around quality in qualitative research (32). As the data were not collected by the research team, respondent validation was not possible. However, we did endeavour to utilise triangulation, clarity, reflexivity, attention to negative cases and fair dealing. Further detail can be found in the Standards for Reporting Qualitative Research form for this work.

### Implications for research and policy

The experiences of the HCWs in our sample suggest the work environment and professional roles have contributed to mental health crises for NHS staff. The experiences of doctors and staff with pre-existing mental health conditions in our sample were particularly poor, a finding which has been echoed in a recent systematic review (31). This suggests these groups are potential targets for workforce interventions, such as the NHS long-term workforce plan (9). However, due to the small number of HCWs samples in this study (n=60), this would need further research to validate. Furthermore, use of the Shout text message service by HCWs in crisis demonstrates the vital importance of the service. It indicates that digital mental health services can overcome traditional support access boundaries, highlighting its benefit during the pandemic and making the case for its ongoing provision.

Policymakers could aim to create tailored packages of support based on risk, for example ensuring HCWs with pre-existing mental health conditions are prioritised on waiting lists for support services. Further the NHS should continue to provide staff with digital mental health services, such as Shout. There is a sense the UK government are “winding down” mental health support for NHS workers post-pandemic, expressed in recent calls for mental health provisions from the *British Medical Journal* and Royal College of Nursing. This collection of personal accounts of crisis demonstrates that digital mental health support provision is vital and needs to continue. Other work on mental health innovations, and workforce recovery post-covid also highlights the hope and opportunity available to restore the NHS workforce with continued personalised, local and co-developed mental health provision(12,33)

It is critical that structures and arrangements are put in place to support the NHS and its workers to be organisationally resilient. This requires significant intervention by policymakers to address the culture of fear and ensure individuals reporting bullying and poor management support are adequately supported. Expectations of management’s duty in pastoral care should be clearly articulated (34,35). Finally, more time and formal networks must be in-built to ensure HCWs are able to build bonds and support networks with their colleagues, particularly given they are increasingly likely to turn to them when in need (34).

## Conclusion

This novel study generated further understanding of the nature of distress and mental health concerns in HCWs. We identified several stressors that may contribute to poor mental health in times of challenging service provision, such as the COVID-19 pandemic, but also in this time of NHS recovery. This qualitative analysis of HCWs’ experiences demonstrates, through powerful personal narratives, that digital mental health support provision is vital and needs to continue.

## Author Statement

Lisa Gould: Idea generation, oversight, data analysis, write up, submission

Eleanor Angwin: Data screening, data analysis, write up

Richard A. Powell: Oversight, write up and submission

Emma L. Lawrance Idea generation, oversight, data collection, write up, submission

## Conflicts of Interest

None

## Funders

Patient Safety Translational Research Centre at Imperial College London. RAP is supported by the National Institute for Health Research (NIHR) Applied Research Collaboration Northwest London. The views expressed are those of RAP and not necessarily those of the NIHR or the Department of Health and Social Care. ELL is supported by Lenore England; without her generosity this work would not be possible.

## Acknowledgements

We would like to thank the team at Mental Health Innovations team, led by Dr Mark Ungless, who gave much time and assistance with the production of this work. We would also like to thank all the healthcare workers who held the NHS together during the COVID-19 pandemic.

## Ethics Approval

The study received ethical approval from Imperial College Research Ethics Committee (19IC5511). All Shout texters were provided information about the use of anonymised data for research purposes and were given the option to opt out of this. The data set was anonymised by Mental Health Innovations and stripped of any personally identifiable information (e.g., names, telephone numbers and addresses) prior to researcher access.

## Data Availability

No data Available. We are unable to share the shout dataset due to the ethical requirements for anonymity for the service users.

## Notes

### Competing Interest Statement

The authors have declared no competing interest.

### Funding Statement

NIHR Northwest london patient safety research

### Author Declarations

The study received ethical approval from Imperial College Research Ethics Committee (19IC5511). All Shout texters were provided information about the use of anonymised data for research purposes and were given the option to opt out of this.

